# The dynamic relationship between COVID-19 cases and SARS-CoV-2 wastewater concentrations across time and space: considerations for model training data sets

**DOI:** 10.1101/2022.11.23.22282684

**Authors:** Rebecca Schill, Kara L. Nelson, Sasha Harris-Lovett, Rose S. Kantor

## Abstract

During the COVID-19 pandemic, wastewater-based surveillance has been used alongside diagnostic testing to monitor infection rates. With the decline in cases reported to public health departments due to at-home testing, wastewater data may serve as the primary input for epidemiological models, but training these models is not straightforward. We explored factors affecting noise and bias in the ratio between wastewater and case data collected in 26 sewersheds in California from October 2020 to March 2022. The strength of the relationship between wastewater and case data appeared dependent on sampling frequency and population size, but was not increased by wastewater normalization to flow rate or case count normalization to testing rates. Additionally, the lead and lag times between wastewater and case data varied over time and space, and the ratio of log-transformed individual cases to wastewater concentrations changed over time. This ratio increased sequentially in the Epsilon/Alpha, Delta, and Omicron BA.1 variant surges of COVID-19 and was also related to the diagnostic testing rate. Based on this analysis, we present a framework of scenarios describing the dynamics of the case to wastewater ratio to aid in data handling decisions for ongoing modeling efforts.

## 1. Introduction

The COVID-19 pandemic stimulated worldwide research on how wastewater-based surveillance of SARS-CoV-2 RNA can be used to monitor infections at the population level. Many studies have found strong correlations between SARS-CoV-2 wastewater RNA samples and COVID-19 cases via diagnostic testing [1–4], and routine wastewater surveillance has supported decision-makers in choosing appropriate public health responses [5–7]. With the widespread availability of at-home tests and decreased severity of disease due to vaccination and/or prior infection, the reliability of reported case data has decreased substantially since December 2021 [8]. To prepare for new surges due to emerging variants or waning immunity there is a need to build forecasting and nowcasting models that use wastewater data as a main input [9,10]. For training, these models require high-quality paired retrospective wastewater and diagnostic testing data. However, both the wastewater and case count data in these retrospective datasets are imperfect, necessitating careful consideration of factors contributing to noise and bias prior to modeling.

### 1.1 Causes of inaccuracies in wastewater data

Concentration of SARS-CoV-2 in wastewater is affected by the number of infected individuals, but also by precipitation events, infiltration and inflow [11], industrial flow contributions, and many other factors [12]. Flow rates at wastewater sampling sites can be used to adjust for dilution, but flow data is not always available, especially for samples collected from manholes or small wastewater treatment facilities where no flow meter is present. Additionally, the heterogeneity of sewage samples and the degradation of SARS-CoV-2 in sewers [13] cannot be accounted for by flow normalization. To address these sources of variability many studies measure cross-assembly phage (crAssphage) or Pepper Mild Mottle Virus (PMMoV) as biological human fecal indicators [3,14,15]. Physicochemical parameters such as total nitrogen, ammonia, conductivity, total suspended solids (TSS), and biological oxygen demand [16,17] can also be used to account for variation in wastewater strength, but they may be substantially affected by industrial inputs [18]). Although the US CDC has published recommendations on the wastewater sampling process and established a reporting database [19], there is currently no overall standard for wastewater SARS-CoV-2 sampling and analysis. Thus, the causes of noise need to be considered individually for each dataset.

### 1.2 Causes of inaccuracies in diagnostic testing data

Diagnostic testing data also includes uncertainty, which may stem from biased allocation of and access to tests across the population, variation in reporting date assigned to each case (e.g. symptom onset, testing date, or date of positive test result), underreporting of at-home test results, and fluctuations in testing rates across time and space [20]. In 2020, the WHO recommended a threshold of 5% test positivity as a metric of sufficient testing. However, this threshold is only valid under certain conditions of contact tracing and sufficient testing of symptomatic individuals, and may only reflect the beginning stages of the pandemic [21]. Generally, case data may be less reliable when testing rates are low, and as of May 26, 2022, Noh & Danuser (2021) estimated a total rate of undetected cases of approximately 55% for California [20]. Modeling testing bias was shown to improve case data accuracy when compared to seroprevalence [22], but normalization in wastewater testing studies is typically focused only on accounting for wastewater strength. Although the importance of assessing testing rates prior to modeling was demonstrated in a recent study [23], to our knowledge, few wastewater studies have directly addressed bias in diagnostic testing data.

### 1.3 Correlation and the ratio between wastewater and case data

Prior research has used correlation between wastewater and case data as a readout for the effectiveness of normalization methods, for determination of lead/lag times between datasets, and as a means to state the value of wastewater monitoring in general [3,4,24–26]. However, statistical caveats of this analysis are often ignored – for example the fact that the correlation of two variables that measure the same phenomenon in a time series is inflated due to autocorrelation [27,28]. Critically, the correlation coefficient reflects the global relationship between the diagnostic testing and wastewater surveillance data and does not offer an insight into the development of this relationship over time. For this purpose, the ratio of log-scaled COVID-19 cases over log-scaled wastewater RNA concentrations may be more appropriate. This ratio should be representative of shedding per person assuming perfect diagnostic testing and accurate wastewater data (not accounting for SARS-CoV-2 RNA decay in the sewer). Log-scaling reduces extreme values in the datasets and mimics a linear relationship between the variables, as they are not normally distributed [7]. Several studies have proposed using this ratio for analysis, and have reported values between 0.24 and 0.39, or up to 0.67 after flow normalization [28–31]. However, time series analysis of this ratio has not been performed on real-world data.

### 1.4 Study objectives

The goal of this study was to investigate the nature of the relationship between wastewater and case data over space and time to provide a basis for future modeling efforts. We present a large, curated dataset with wastewater and case data collected in California during the first two years of the COVID-19 pandemic, when case data quality was high. Our analyses reveal the instability of the relationship between wastewater and case counts and identify three main variables that could affect models for predicting cases from wastewater: dynamic lead/lag, changes in fecal shedding due to viral variants, and changes in reporting of individual cases to public health departments.

## 2. Materials & Methods

### 2.1 Wastewater sample collection and analysis

Raw wastewater samples (n=2480) were collected via 24-hour flow-or time-weighted composite samplers from 26 sewersheds in California between 1 to 5 times per week (**Table S1, Table S2**). All sewer systems had separate storm sewers, with the exception of system D, where wastewater and storm sewers were combined. Sampling dates ranged between October 2, 2020 and June 29, 2022, although not all sewersheds were sampled for the full time period. Sample collection points were at wastewater treatment plant influent (“sewersheds”) and at pump stations and manholes (“sub-sewersheds”). Samples were aliquoted (40 mL) into tubes containing the 4S method lysis mixture, shipped overnight to UC Berkeley, and analyzed according to the laboratory procedure described by Kantor et al. [32]. Analysis used the 4S method for total RNA extraction [33] followed by RT-qPCR for SARS-CoV-2 CDC N1, Pepper Mild Mottle Virus, and Bovine Coronavirus [3]. Quality controls included extraction negative controls, duplicate extractions, extraction spike-in controls (Bovine Coronavirus), triplicate RT-qPCR reactions, no-template controls, and standard curves, as described in Kantor et al. [32]. Data not passing quality control were removed and were replaced with repeated analyses wherever possible.

### 2.2 Wastewater data preparation

Wastewater data were preprocessed as previously described [32]. Briefly, RT-qPCR outliers were removed, Cq values were converted to gene copy numbers using an aggregated standard curve, RT-qPCR replicates were combined by taking the geometric mean, and sample weight was used to calculate the gene copies per milliliter of wastewater. Extraction replicates were combined by taking the geometric mean. Five outliers that could be directly attributed to changes in plant operations or autosampler failures were manually removed.

The wastewater concentration was normalized by flow to reduce the effects of dilution by precipitation, groundwater infiltration, and industrial wastewater. Precipitation data for the years 2020-2022 were downloaded from the NOAA (National Oceanic and Atmospheric Administration) website for each county [34]. Using this dataset, we calculated the median dry flow for each sewershed by taking the median of daily flow rates for all days that were recorded as dry (precipitation < 0.2 inches) within the county. We then used this median dry flow to recalculate the SARS-CoV-2 RNA concentration in the wastewater and removed the industrial proportion of flow estimated by the wastewater agencies from the daily flow, as in Eq. 1 (Method 1). A second variation on this method (Method 2) entailed normalizing values only for days on which precipitation occurred. For Method 2, values were normalized according to Eq. 1 to offset a potential dilution and remove the industrial flow proportion, and all other values were normalized according to Eq. 2 to remove only the industrial flow proportion. We tested different time frames of up to three days after rain events to account for potential delays in the effect of precipitation on the dilution of the signal, however, including only the day of the rain event resulted in the highest correlations (not shown).

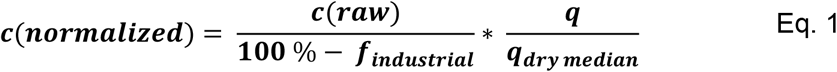

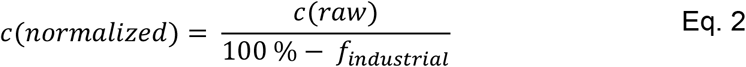

Where c(normalized) is the flow-normalized RNA concentration (gc/mL), c(raw) is the measured SARS-CoV-2 RNA concentration (gc/mL), q is the daily flow (MGD), q_dry median_ is the median dry flow (MGD), f_industrial_ is the percentage of total flow estimated to come from industrial sources.

Normalization with PMMoV, TSS and conductivity was performed according to the following Eq. 3.

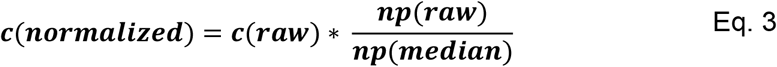

Where c(normalized) is the normalized SARS-CoV-2 RNA concentration (gc/mL), c(raw) is the measured SARS-CoV-2 RNA concentration (gc/mL), np(raw) is the concentration of the normalization parameter, and np(median) is the median concentration of the normalization parameter.

Previous studies recommend applying a 7-day or 10-day moving average to the wastewater data [23]. However, as the sampling frequencies in our dataset varied over time and space, lowess smoothing and interpolation was chosen for analysis of lag and lead times and for data visualization [3]. The smoothing coefficient alpha was defined as alpha = X/n, where n was the total number of data points at a given site. We note that because lowess smoothing depends on the total number and density of data points, it may have led to slightly different effects on data from different sites. Unless stated, other analyses were performed on the original wastewater dataset to maintain the integrity of the recorded data.

### 2.3 COVID-19 case data collection and preparation

Masked daily case counts per sewershed were provided by the California Department of Public Health, based on sewershed boundaries provided by wastewater agencies. Cases were attributed to the earlier of 1) the date of diagnostic testing or 2) the reported date of first symptoms, when both dates were available. Sewershed population estimates were based on reports by the wastewater agencies and, if unavailable, government census data (**Table S1**). Daily case counts were masked below 3 cases for sewersheds representing populations of 200,000 or less, and below 5 cases for populations of 50,000 or less, but instances of zero cases were reported as zero. During data preparation, masked values were filled with the mean of the masked ranges **(Table S3)**. Case counts and testing rates were normalized to a population of 100,000 and a centered 7-day moving average value was calculated to smooth weekly periodicity. For log-scaled analyses, days with zero average cases were dropped prior to analysis.

### 2.4 Normalization of case data to account for diagnostic testing rates

County-level diagnostic testing rate data were acquired from publicly available sources [35]. In order to compensate for fluctuations in how accurately the case count data reflected the true incidence of infection, we adjusted the reported cases to the diagnostic testing rates according to the following equations. Equation 4 linearly inflates the daily cases according to the fraction of utilized testing capacity on a given day. Equation 5 compensates for a positivity rate bias as defined by Chiu and Ndeffo-Mbah, 2021 [22].

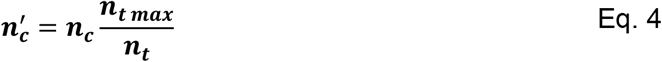

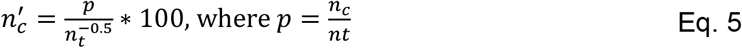

Where n_c_’ is the adjusted number of cases, n_c_ is the original number of cases, n_t_ is the number of tests, n_t max_ is the maximum number of tests ever reported on a single day within the study period, and p is the test positivity rate. As described in Section 2.3, all values are normalized by sewershed population size.

### 2.5 Data analysis

The data analysis pipeline was created in Python 3.7 using the Pandas v1.3.5 and Numpy v1.21.6 libraries. The rank correlation of smoothed daily cases with raw and normalized wastewater sampling data was quantified for each sewershed using the Kendall’s Tau b coefficient (SciPy v1.7.3) [3]. Autocorrelation and lowess smoothing were calculated using Statsmodels v0.10.2, and data visualization was performed using Plotnine v0.9.0.

For analyses of individual surges, the following timeframes were used: the first major surge we observed (including Epsilon, Alpha, and other minor variants) was defined from the start of the time series (October 2020) to April 15, 2021, the Delta surge from April 16 to November 26, 2021, and the Omicron BA.1 surge from November 27 to March 15, 2022 based on California Department of Public Health [36] and COVID-CG [37] and our wastewater sequencing data (unpublished).

To assess the stability of the relationship of log-scaled COVID-19 cases and log-scaled wastewater SARS-CoV-2 concentrations, we implemented a linear regression model with Scikit-Learn v1.13 using these inputs during the Epsilon/Alpha variant surge for sewersheds D1, D2, K, L, and M. This model was then applied to the subsequent Delta and Omicron variant surges and evaluated using the R^2^ goodness-of-fit parameter (**Table S4**). The data analysis pipeline and all necessary datasets are available at GitHub (github.com/RebeccaSchill/WBE).

## 3. Results and Discussion

We analyzed the SARS-CoV-2 RNA concentration in 2480 wastewater samples from 26 sewersheds sampled between 1-5 times per week from approximately October 2020 - April 2022. This data was paired with sewershed-specific COVID-19 daily case counts and county-level diagnostic testing rates and positivity rates. Populations of the sewersheds ranged from 12,000 to 4 million, and flow rates ranged from 0.2 to 243 million gallons per day (**Table S1**). Precipitation was infrequent (0% - 26% of days in the time series for each site), due to a combination of drought and mediterranean climate in California.

### 3.1 Denoising via normalization of wastewater and case data

We first compared methods for removing noise from the wastewater and case data. As previously described, denoising efficacy was evaluated based on changes to Kendall’s tau calculated for the relationship between wastewater and case data (**Table 1**) [3,38]. Flow normalization marginally improved the correlation for 14 sewersheds, but the effect of normalization was minimal, likely because of infrequent precipitation (**Table S5**). Normalization of the wastewater data from two major sewersheds (D1 and D2) to PMMoV, TSS, or conductivity also did not increase correlations with case data (**Table 1, Table S6**). Other studies have shown that normalization of wastewater to PMMoV can decrease noise, but successes have been inconsistent and appear to be dependent on the laboratory method used for virus concentration and extraction, as well as sewershed size [39–41], and possible dietary variation. Our laboratory method for RNA extraction (4S, Whitney et al., 2021) lacked bead-beating and therefore may not have achieved complete and consistent lysis of PMMoV, required for accurate quantification.

**Table 1.** Average Kendall’s correlation of log-scaled values (for all chosen sewersheds) before and after applying different normalization methods to 7-day moving average case data and unsmoothed wastewater data. Averages represent all 26 sewersheds.

|  | Wastewater data normalization methods |  |  |  |
| --- | --- | --- | --- | --- |
| Case data normalization methods | No normalization | Flow (Method 1, Eq. 1) | Flow (Method 2, Eqs. 1 & 2) | PMMoV |
| No normalization | 0.57 | 0.58 | 0.57 | 0.47 |
| Normalized by testing capacity (Eq. 4) | 0.50 | 0.50 | 0.49 | 0.12 |
| Normalized by testing bias (Eq. 5) | 0.54 | 0.54 | 0.53 | 0.14 |

Normalization of the sewershed-level case counts to the county-level diagnostic testing rate (Eq. 4) reduced the strength of the correlation to wastewater data. Accounting for the test positivity rate in addition to testing rate in a bias function (Eq. 5) resulted in a more modest decrease in correlation (**Table 1**). This suggests that additional calibration of the testing bias model (e.g. with regional seroprevalence data) is likely required.

### 3.2 The correlations between wastewater and case data differed by sewershed

Flow-normalized Kendall’s tau for wastewater and case data from different sewersheds exhibited a wide range, from 0.27 to 0.74 (**Figure 1**). In general, larger treatment plants with more frequent sampling and less masking of individual case data showed the highest tau values (**Table S3**). Meanwhile, smaller sewersheds appeared subject to higher noise, for several possible reasons. First, when the total absolute number of infected individuals are low, as is often typical in small sewersheds, each individual contributes a higher fraction of the total wastewater SARS-CoV-2 concentration, and sampling effects (e.g. missing a flush) can create more noise [12]. Additionally, the effect of mobility (e.g. one infected person entering or leaving the sewershed) is stronger [42]. Second, consistent with recommendations from the US CDC [43], we found that sewersheds with fewer than 2 samples per week tended to produce weaker correlations, and these were often smaller treatment facilities. This is in line with reports of lower sampling capacities at smaller wastewater treatment plants [44]. Many of these small sewersheds also had fewer than 50 total sampling events (**Table S1**). Additionally, we note that the time series of wastewater and case counts were autocorrelated (Durbin-Watson statistic d < 1.5 in all sewersheds), and autocorrelation may have increased with increasing sampling frequency, affecting tau values differently in each sewershed.

**Figure 1.**
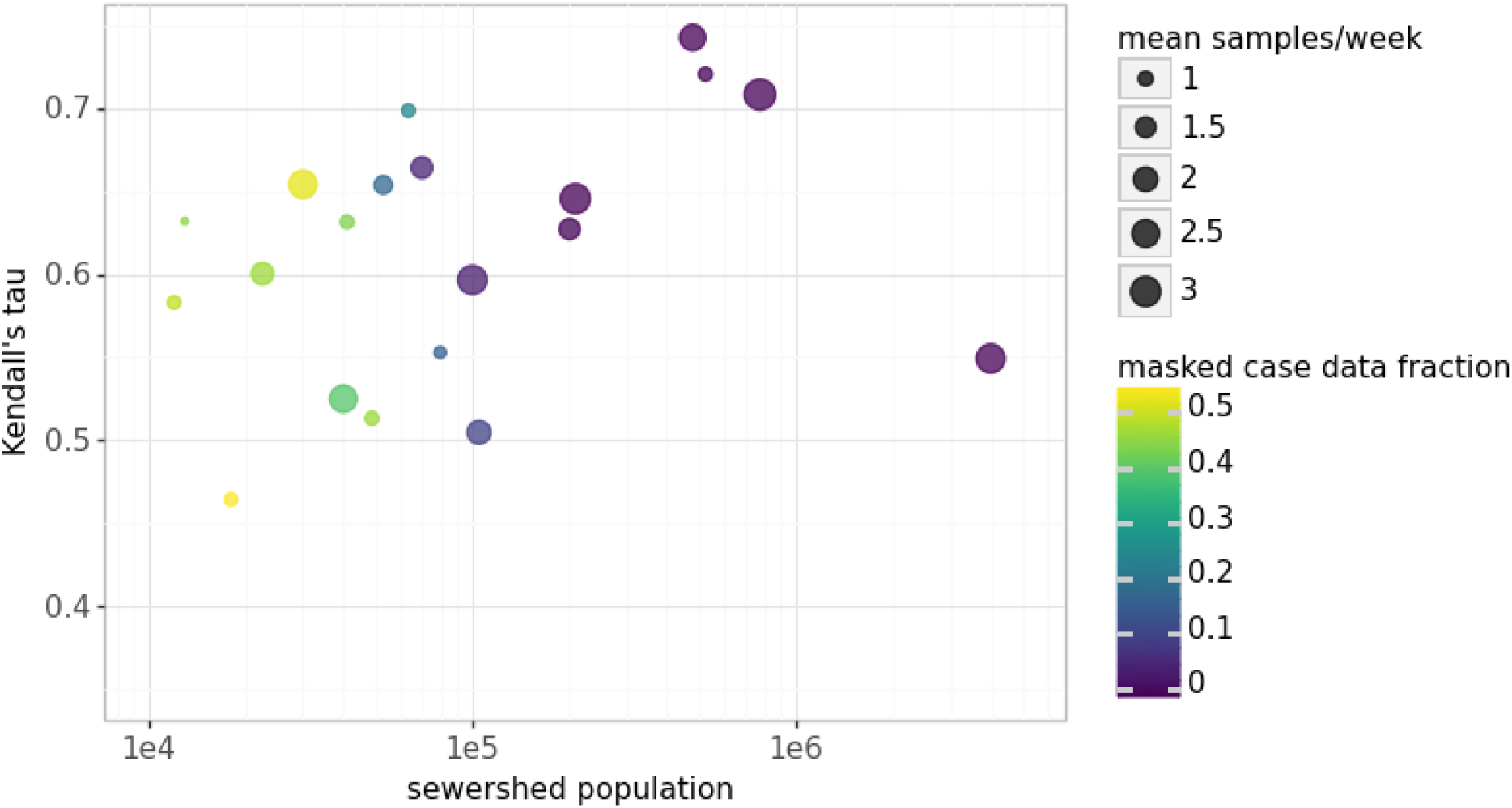
The Kendall’s correlation coefficient between log-scaled cases and unsmoothed log-scaled wastewater SARS-CoV-2 RNA concentrations (y-axis) increased with increasing sewershed population (x-axis) and was affected by weekly sampling frequency (point size) and by the fraction of case data that was masked (point color).

Third, a larger proportion of daily case data was masked in the smallest sewersheds, with a median value of 45% of data masked (**Table S3**). Two sewersheds with a masking proportion >95% were removed from further analyses. Lastly, within-sewershed fluctuations in diagnostic testing rates may also have led to differences in wastewater-case correlation between sewersheds [45] (see Section 3.6). We were unable to assess disparities in testing rates given that testing rate data were available at the county level only, which may not be representative of individual sewersheds. Overall, the collection of high-resolution datasets improves the reliability of the relationship between case counts and wastewater data and the accuracy of forecasting models [46]. These findings motivate policy to report detailed diagnostic testing and COVID-19 case data and to provide support for smaller communities to increase wastewater sampling frequency in locations where case data may be the least accurate [47].

### 3.3 Lag between case data and wastewater data was dynamic over time and space

Modeling work may need to take into consideration the lead/lag between case counts and wastewater data. As previous studies have reported wastewater lead times over case data of between 0 and 14 days [48], we hypothesized that lead time could vary substantially by sewershed and over time due to factors such as evolving virus variants, sewage travel distance, and access to and frequency of diagnostic testing [49]. To assess lead/lag times, we first smoothed the wastewater data to remove noise (see Methods; Section 2.2), then calculated the cross-correlation Kendall’s tau-b between the flow-normalized wastewater data and case data shifted in each direction by 1 to 14 days (see Methods; **Figure S1A**). A wastewater lag/lead time between -3 days and +4 days was detected in four of the seven sewersheds that were sampled throughout the entire time series, but the corresponding increases in Kendall’s tau-b were very low with a maximum increase of 3%.

We next examined whether the wastewater lead/lag changed during periods when different variants predominated. Overall, varying wastewater lead times from +1 to +13 days were observed in 12 of 17 sewersheds during the Epsilon/Alpha variant-dominated surge. This lead time was also observed in 14 out of 20 sewersheds during the Delta variant surge but faded during the Omicron variant surge, where wastewater data lagged and led case data in an equal number of sewersheds (**Figs. S1B, S1C, S1D**). The dynamic behavior of the time shift between wastewater and case data across variants is demonstrated in detail at two sewersheds (D1 and K) in **Figure 2**. Notably, during the first surge we observed, the peaks in wastewater and case data are not aligned, resulting in very long lead times. This is likely due to a combination of testing fluctuations over the winter holidays and the multiple overlapping surges of different variants (Epsilon, Alpha, Gamma, and others). The wastewater lead time lessened significantly during the Delta surge in both sewersheds and disappeared during the Omicron surge. This aligns with previous reports of reduced wastewater lead times after the Alpha surge [7,50].

**Figure 2.**
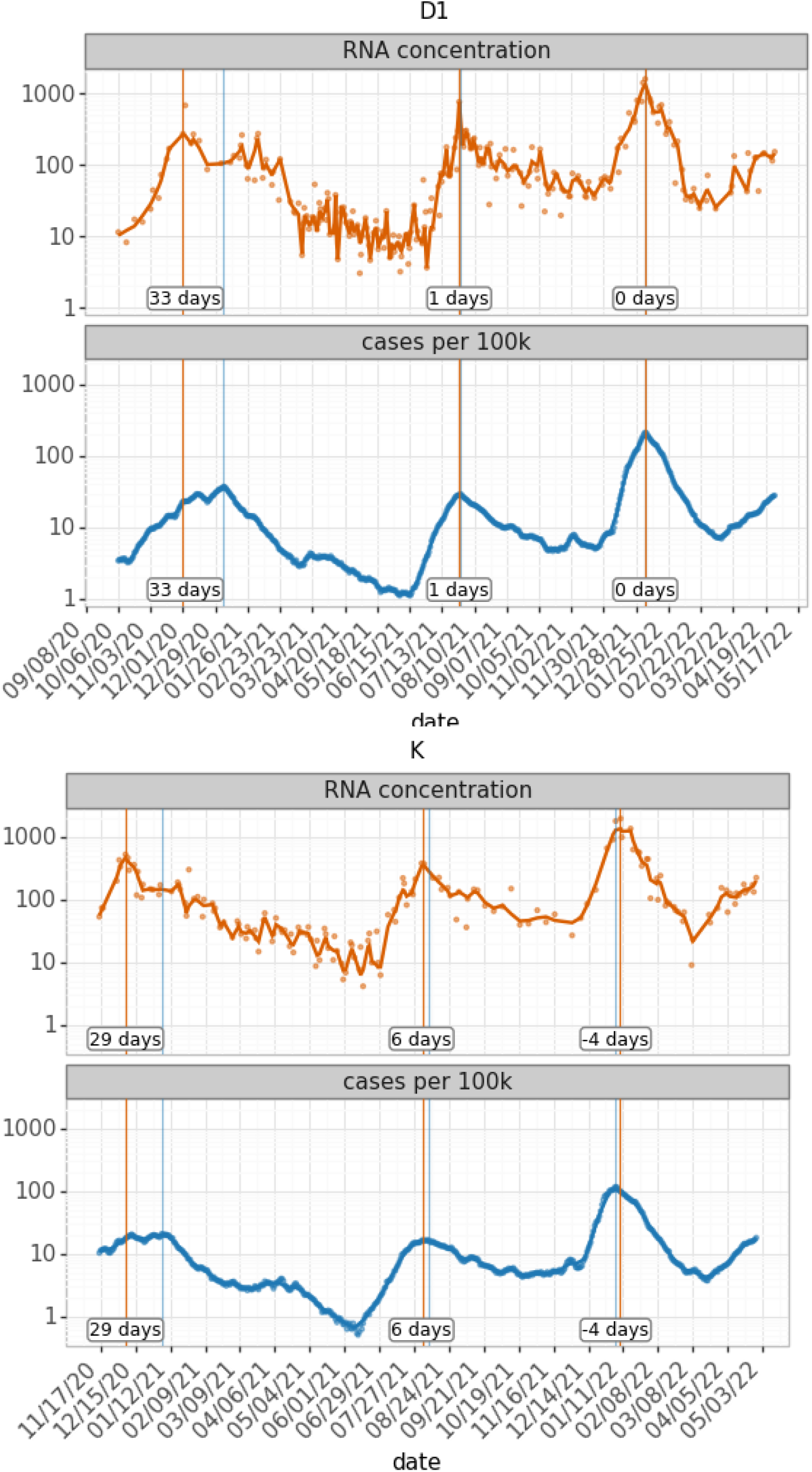
Lag between wastewater and individual case data varied over time and between sewersheds. Flow-adjusted wastewater SARS-CoV-2 concentrations in gene copies per milliliter (orange), COVID-19 cases per 100,000 people (blue), including a lowess smoother (alpha=0.05) are shown for two sewersheds. Sewershed D1 (top) represents a population of 750,000, with high sampling frequency, while sewershed K (bottom) represents 480,000 people with intermittently reduced sampling frequency. Vertical lines indicate minima and maxima of the timeseries of cases (blue) and wastewater (orange). Labels indicate the wastewater lead time at the surge peak in days.

The use of cross-correlation to determine lag/lead times between case data and wastewater data is based on the assumption that there is a static lag between the two datasets. Static lag could reasonably stem from near-constant factors such as sewer transit time (constant within a sewershed) or delay between infection and symptom onset that would trigger diagnostic testing (assumed constant for each variant). However, our findings of dynamic lag over time and across sewersheds suggest that other factors are at play. Wastewater sampling frequency, population-level immunity, or changes in diagnostic testing strategy/availability differed between surges and locations and likely affected lead times. Previous studies have highlighted that lead time calculations need to be adapted to different purposes, for example real-time decision-making versus retrospective data analysis [48]. In this study, due to the applied smoothing methods, lag calculations do not represent real-time data availability, but instead reveal a potential delay in measurable signal between wastewater and diagnostic testing. Our findings of dynamic lead times suggest that cross-correlation, and by extension, simple linear regression models (**Table S4**), are therefore insufficient for describing the relationship between case and wastewater data for retrospective data analysis, and dynamic lead times will affect input data for modeling.

### 3.4 The ratio of cases per wastewater RNA was not constant over time and space

To explore the dynamic nature of the relationship between wastewater and case data, we calculated the ratio of log(cases) per log(wastewater concentration) (see **Figure S2** for example). For five large sewersheds sampled continuously throughout the analyzed time frame (**Figure 3**), we found that the magnitude of the ratio was different at each sewershed, likely affected by the accuracy of the population estimates. The ratio also changed over time: during the Epsilon/Alpha surge, the ratio remained stable overall before decreasing to a minimum just before the peak of the Delta surge. The ratio then recovered and increased to a maximum during the first Omicron surge. Towards the end of this surge, the ratio decreased once more. These developments were similar at sewersheds D1, D2, and K, but less pronounced or more stochastic in sewersheds L and M and others where sampling was less frequent (**Figure 3;** see **Figure S3** for all sewersheds).

**Figure 3.**
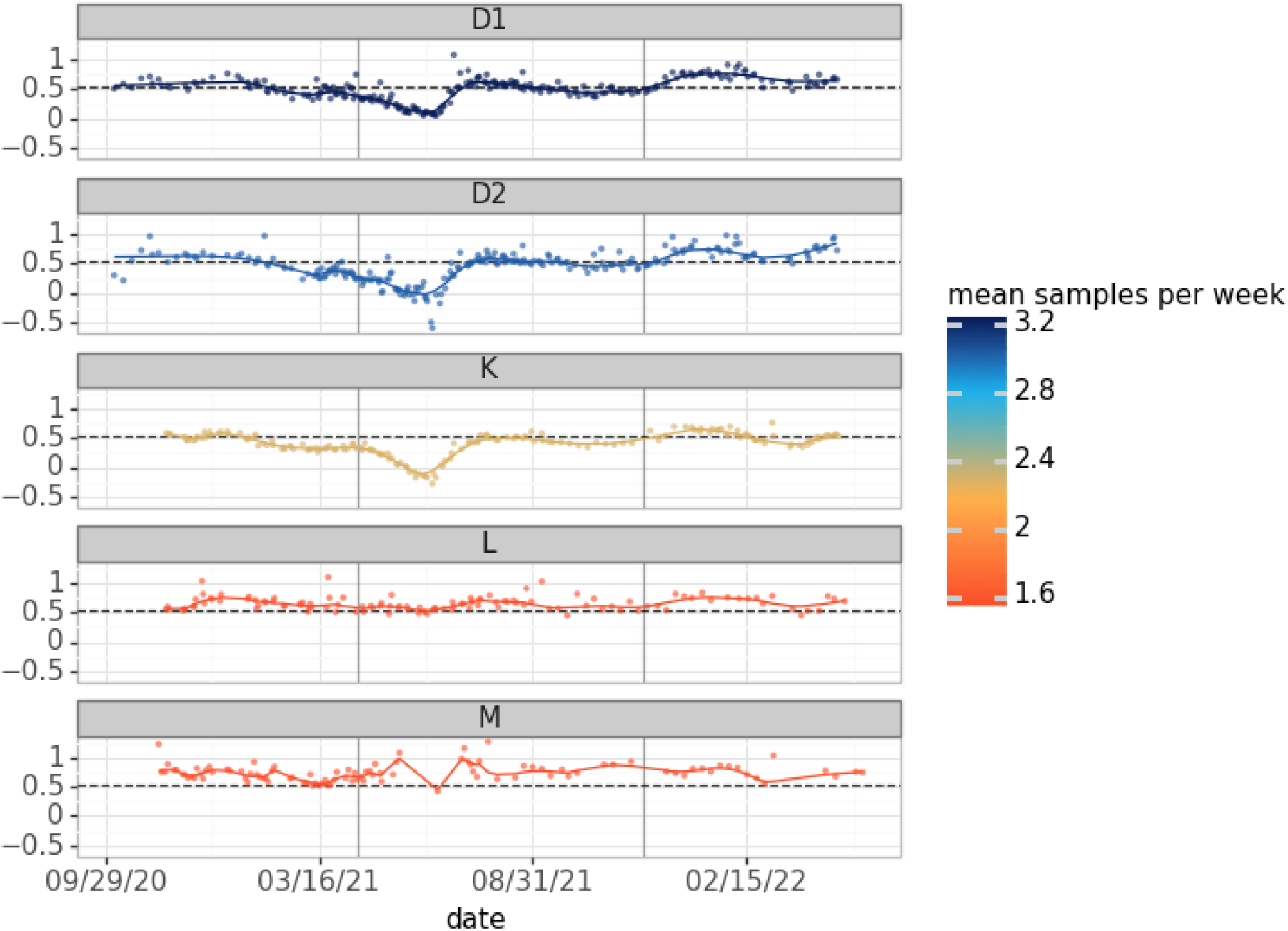
Ratio of log(cases per 100,000) over log(wastewater concentration) at sewersheds D1 (n=262), D2 (n=244), K (n=172), L (n=121), and M (n=131), for three surges (separated by vertical gray lines). Smoothed lines were generated with lowess (alpha=0.05), and the dashed line represents the median ratio across all 5 sewersheds (0.55). For each sewershed shown, case data masking was below 5% of all data points. Two outliers were removed at sewershed M for visualization.

### 3.5 Ratio of cases per wastewater RNA differed by variant

To test for the effect of evolving virus variants on wastewater surveillance data, we calculated point estimates for the ratios in each sewershed as follows: for each variant surge, we identified the maximum number of cases per 100,000 people (centered 7-day average) and the maximum lowess-smoothed wastewater concentration reported. Then, we calculated the ratio by dividing the log-scaled maximum cases by the log-scaled maximum wastewater concentration. We could not isolate the Epsilon and Alpha variants, as the surges partially coincided. Although our analysis was limited to 5 sewersheds, we observed a significant increasing trend in the ratio from the Delta variant to the Omicron variant (**Figure 4**). The decrease between the Epsilon/Alpha and Delta variants could be observed as well but the difference was not statistically significant. The ratio appears consistently lower for sewershed K but sewershed-specific differences were not significant (Kruskall-Wallis, p = 0.429)

**Figure 4.**
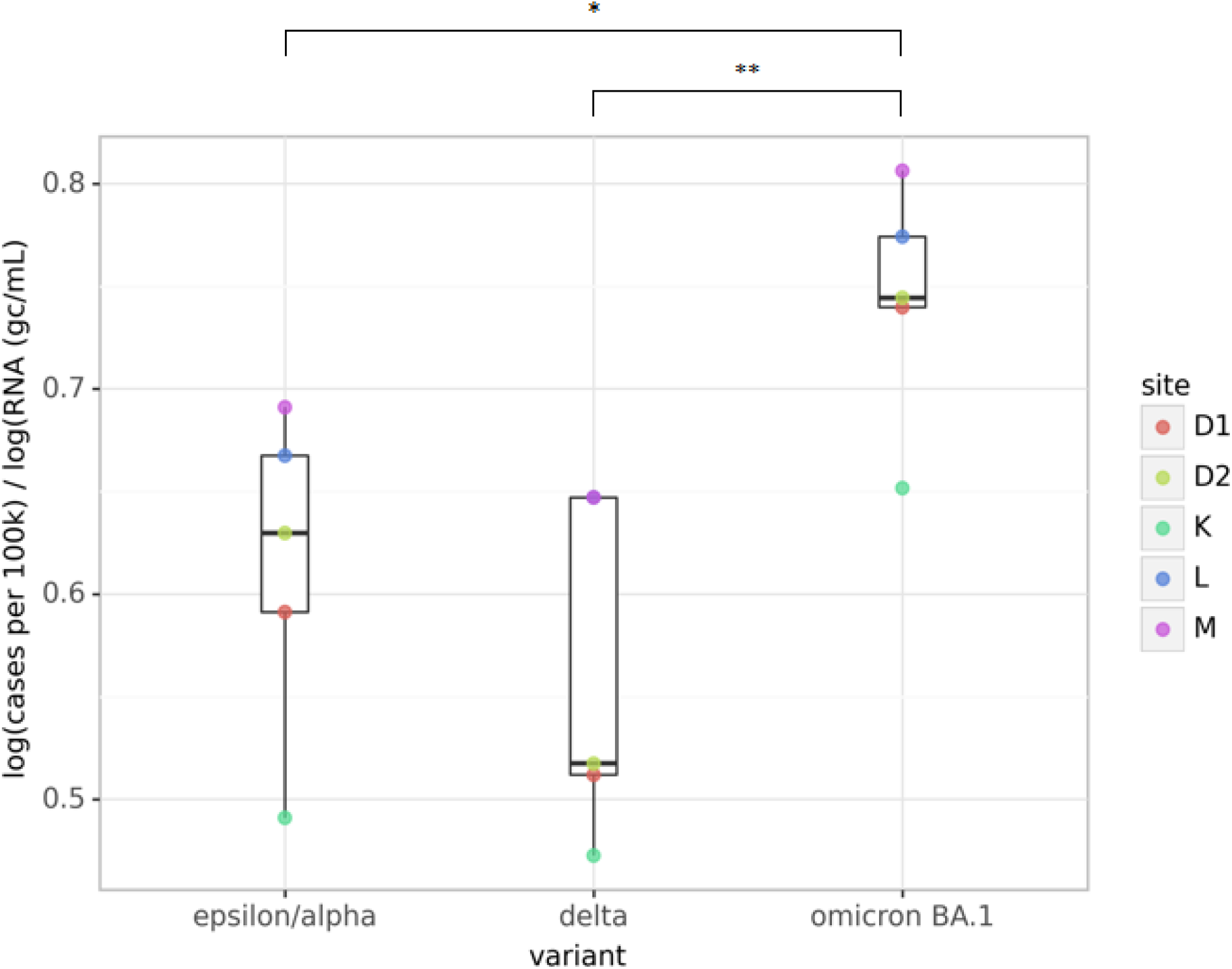
The ratio of log(cases) over log(wastewater RNA) changes for each variant at sewersheds D1, D2, K, L, and M. The ratio was calculated from the peaks of the three surges after lowess smoothing (alpha = 5 / total samples). The difference between the Omicron BA.1 variant and the other variants was statistically significant (Mann-Whitney, * p = 0.032, ** p = 0.008), while the difference between the Epsilon/Alpha and other variants to the Delta variant was not (p = 0.421).

Additionally, we found that a linear regression model trained to predict case data from flow-normalized unsmoothed wastewater data for Alpha/Epsilon surge deteriorated in fit during the Delta and Omicron BA.1 variants (see **Table S4**). Overall, these findings agree with the reports of increased fecal and oro-nasopharyngeal viral loads during the Delta surge [51,52] and with reduced fecal shedding observed with the Omicron variant [53,54]. While SARS-CoV-2 oro-nasopharyngeal viral load was reportedly reduced after vaccination [55–57], more research is needed to determine potential effects of vaccination and prior infection on fecal shedding rates.

### 3.6 Diagnostic testing rates influenced the cases-to-wastewater RNA ratio

Given the drop in the cases-to-wastewater RNA ratio during the pre-Delta period, we hypothesized that changes in diagnostic testing dynamics might influence this ratio. Indeed, we observed that low testing rates corresponded with low ratios throughout the time series, including during the pre-Delta trough (**Figure 5**). Correlations between cases-to-wastewater ratios and diagnostic testing rates over time were significant in several sewersheds (**Table S7**). Notably, the strengths of these relationships differed for sewersheds in the same county (**Table S7**), suggesting that sewershed-level testing rates differed from those at the county level or the quality of wastewater data differed for sewersheds in the same county. Additionally, the measurement uncertainty was likely higher during periods of low case counts and low wastewater concentrations, which could also have contributed to the change in the ratio observed during these periods.

**Figure 5.**
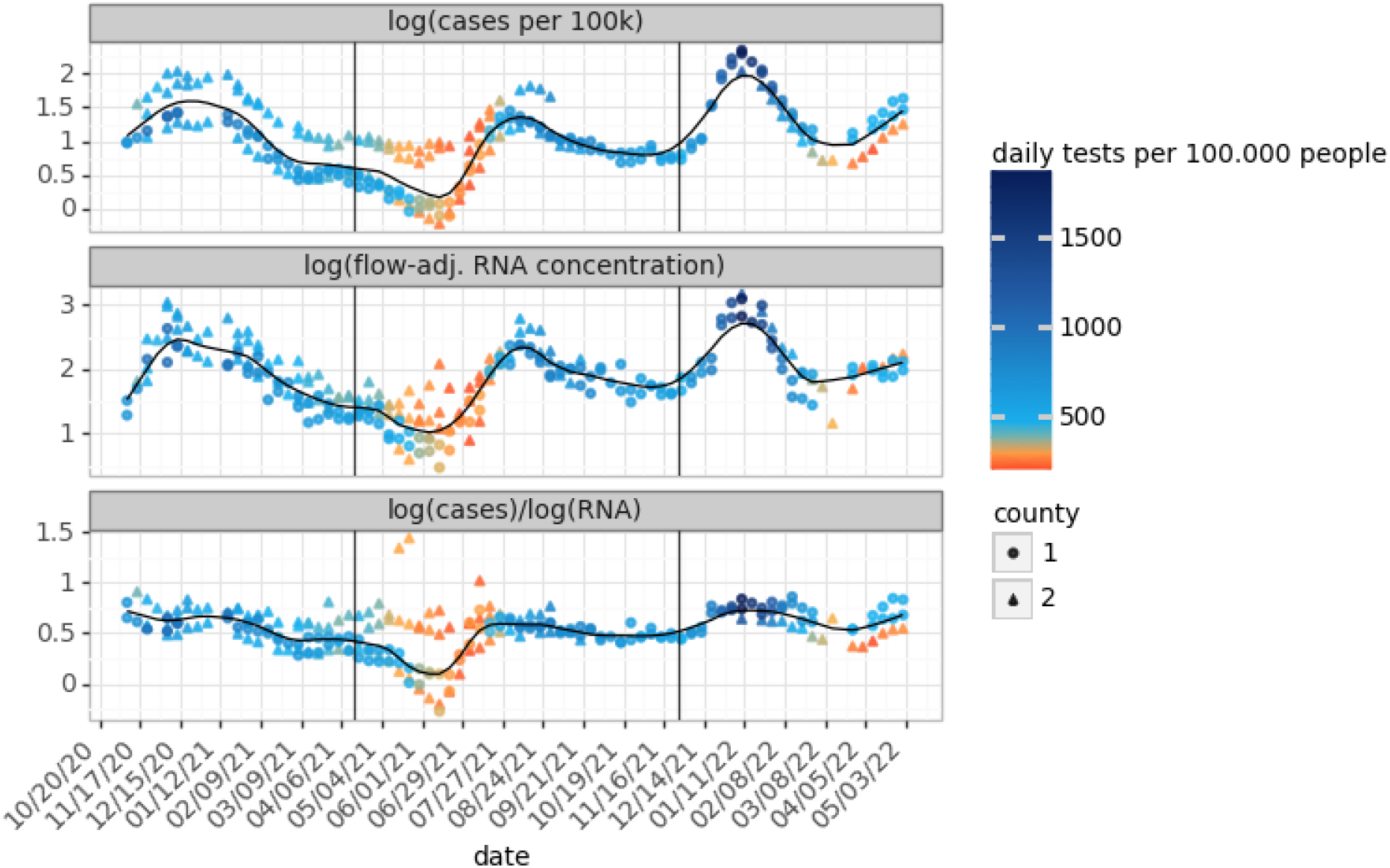
Time series from 5 sewersheds (D1, D2, K, L, M) of the median weekly COVID-19 cases per day (top), median weekly flow-adjusted wastewater SARS-CoV-2 RNA (gc/mL) (middle) and the ratio between them (bottom). For each sewershed, for a given week, a minimum of two data points was required for a weekly median to be shown. The color of the data points represents the daily diagnostic testing rate per 100,000 people, and the shape indicates the county in which the sewershed is located.

Although low diagnostic testing rates may partially explain low ratios between surges, we note that the cases-to-wastewater RNA ratio recovers more quickly than the testing rates, suggesting that undertesting cannot be the only cause of lower ratios (**Figure 5**). This is underlined by the fact that normalizing by testing rates (via Eq. 4 and 5) did not completely flatten the cases-to-wastewater ratio over time (**Figure S4**). Critically, differences in the slopes of the case and wastewater curves may also have affected the ratio between them. As has been shown in previous studies [23], we suggest that proportionally more cases remained undetected at the very beginning of a surge until the diagnostic testing rates adapted, as the case curves increased more steeply than the wastewater curves before the peak of each surge (**Figure S5**). This affected the ratio as well (**Table 2**). After the peak of each surge, the decline in wastewater RNA concentrations was more gradual than the decline in cases, perhaps due to prolonged fecal shedding [58]. We echo the suggestion by Daza-Torres (2022), that input data for modeling should be drawn from time periods with adequate testing. Future work could assess testing behavior and the distribution of tests across the population (e.g. symptomatic vs. asymptomatic, retesting, etc.), to further adjust case data.

**Table 2.** Scenarios that can lead to changes in the ratio of log transformed case to wastewater data.

| Ratio | Changes to factors contributing to case data | Changes to factors contributing to wastewater signal | Both |
| --- | --- | --- | --- |
| **Increase** | Increased diagnostic testing rates without proportional increase in incidence | Increase in incidence that is captured in the case data but is not reflected in wastewater data due to lower shedding rate or duration | Tested cases increase before this is reflected in the wastewater data<br>Tested cases increase more steeply than is reflected in the wastewater data |
| **Decrease** | Increase in incidence reflected in wastewater data is not reported in case data due to undertesting | Higher wastewater values due to increased shedding rate or duration | Wastewater values increase before cases reflect the change (undertesting) |

## 4. Conclusions

Based on our observations, we define a framework of key factors that may affect the cases-to-wastewater SARS-CoV-2 RNA ratio over time, encompassing variation in diagnostic testing rates, changes to fecal shedding, and fluctuations in the temporal off-set between wastewater surveillance and case count data (**Table 2**). Future efforts could model these factors to come to a more accurate understanding of the ground truth case counts. Additional work could also incorporate hospitalization [7], vaccination, mobility and other data types that were not considered here. Importantly, modeling work should ensure that the case data used to train a predictive model are drawn from a period(s) when testing was adequate [23]. Additionally, we found that within our dataset, wastewater data varied in quality, and our analysis was limited by the changing frequency of wastewater sample collection throughout each time series. Thus, wastewater data to be used for modeling requires careful curation and potentially smoothing. However, we observed that smoothing led to the loss of extreme values, many of which were important maxima and minima. Smoothing can hide or delay rapid changes in the time series, affecting the lead/lag between wastewater data and case counts. Future work should compare raw and smoothed model inputs to ensure that smoothing maintains the integrity of the data.

Looking forward, once sewershed-specific models have been established, subsequent modeling will benefit from the fact that the sewersheds themselves will remain relatively consistent: the structure of the sewer system itself, transit time of sewage, noise from precipitation, and industrial discharge can be taken into account with ongoing data and existing normalization methods. These models will be independent from case counts and testing, and thus independent from lead/lag times relative to cases. The key factor subject to change will be fecal shedding rate and duration (**Table 2**). Models will require updated *in vivo* studies of fecal shedding profiles to adjust for new SARS-CoV-2 variants and evolving immunity in the population.

## Data Availability

All wastewater data are available in the Supplementary Materials. For privacy reasons, COVID-19 case data used in this study are not shared and may be requested directly from the California Department of Public Health.

https://github.com/RebeccaSchill/WBE

## Supplementary Materials

**Figure S1A**. Kendall’s correlations between wastewater SARS-CoV-2 RNA and COVID-19 cases per 100,000 people without timeshift (left) and cross-correlation heatmap of changes in Kendall’s tau after shifts of up to -14 to +14 days were applied to case data (right). The maximum correlation is indicated by gray points. Prior to correlation calculations, all wastewater data were flow-normalized, log-scaled, and lowess smoothed (with interpolation) to reduce noise, and all case data were converted to 7-day moving averages and log-scaled. Sewersheds were included only if they were sampled throughout the entire time series.

**Figure S1B**. Kendall’s correlations between wastewater SARS-CoV-2 RNA and COVID-19 cases per 100,000 people during the Epsilon/Alpha variant-dominated surge without time shift (left) and after shifts of up to -14 to +14 days were applied to case data, (right), as in Figure S1A.

**Figure S1C**. Kendall’s correlations between wastewater SARS-CoV-2 RNA and COVID-19 cases per 100,000 people during the Delta variant-dominated surge without time shift (left) and after shifts of up to -14 to +14 days were applied to case data, (right), as in Figure S1A.

**Figure S1D**. Kendall’s correlations between wastewater SARS-CoV-2 RNA and COVID-19 cases per 100,000 people during the Omicron BA.1 variant-dominated surge without time shift (left) and after shifts of up to -14 to +14 days were applied to case data, (right), as in Figure S1A.

**Figure S2**. Time series of flow-normalized wastewater SARS-CoV-2 RNA concentration (gc/mL, orange), 7-day moving average COVID-19 cases per 100,000 people (blue), the ratio of log-scaled cases over log-scaled RNA concentration (gray), and lowess-smoothed curves (alpha = 0.05), at site D1 from August 25, 2020 to May 31, 2022.

**Figure S3**. Time series of the ratio between log10(COVID-19 cases) and log10(wastewater SARS-CoV-2 RNA concentration) at 24 sites. Graphs are sorted by sewershed population and colored by mean weekly sampling frequency. Fraction of case data that is masked is represented by the following: *0-0.05 **0.05-0.33 ***>0.33. One outlier at site F was removed for visualization.

**Figure S4**. Lowess-smoothed (alpha=0.1) ratio of log(cases) to log(wastewater concentration) non-normalized, normalized by testing capacity (Eq. 4) and normalized by testing bias (Eq. 5) at 24 sewersheds. Averages were calculated for each week and each sewershed. For each sewershed, for a given week, a minimum of two data points was required for inclusion.

**Figure S5**. Lowess smoothed time series of log-scaled, flow-normalized wastewater SARS-CoV-2 RNA concentration (gc/mL, orange) and log-scaled, 7-day moving average COVID-19 cases per 100,000 people (blue) at sites D1, D2, K, L, and M. Both time series were rescaled by defining the minimum as 0 and the maximum as 1.

**Table S1**. Overview of sites and sampling information.

**Table S2**. Input wastewater dataset.

**Table S3**. Summary of sewersheds grouped by serviced population.

**Table S4**. R2 values of linear regression trained on the relationship of log-scaled COVID-19 cases and log-scaled wastewater SARS-CoV-2 RNA concentration during Alpha/Epsilon variant surge in each sewershed.

**Table S5**. Differences in Kendall’s Tau-b correlation coefficient after flow normalization using two methods (see Section 2.2), in relation to the coefficient of variation of flow at the respectives sites.

**Table S6**. Kendall’s tau values for the correlation between log10(cases per 100,000) and log10(wastewater SARS-CoV-2 RNA concentrations) with normalization.

**Table S7**. Kendall’s correlation coefficient for the relationship between sewershed cases-to-wastewater RNA and county-level testing rate.

## Acknowledgements

We thank the members of the UC Berkeley wastewater testing laboratory, including Joaquin Bradley Silva, Christina Lang, Matt Metzger, Melissa Thornton, student laboratory assistants, and volunteers. We are deeply grateful to our partners at wastewater agencies in California for sample collection and provision of wastewater treatment plant operations data. We acknowledge CDPH for providing sewershed-bounded case data and county-level testing rate/positivity rate data and for helpful discussions during the data collection phase of this work.

## Author Contributions

Conceptualization, RSK, KLN, and SHL; Methodology, RS, RSK; Formal Analysis, RS; Data Curation, RS, RSK; Writing – Original Draft Preparation, RS, RSK; Writing – Review & Editing, RSK, KLN, and SHL; Visualization, RS; Supervision, RSK, KLN; Project Administration, RSK; Funding Acquisition, RSK, KLN, and SHL. All authors read and approved the final manuscript.

## Institutional Review Board Statement

Ethical review and approval were waived for this study, due to the anonymized, aggregated nature of the data.

## References

1. Bonanno Ferraro, G.;Veneri, C.;Mancini, P.;Iaconelli, M.;Suffredini, E.;Bonadonna, L.;Lucentini, L.;Bowo-Ngandji, A.;Kengne-Nde, C.;Mbaga, D.S.;et al. A State-of-the-Art Scoping Review on SARS-CoV-2 in Sewage Focusing on the Potential of Wastewater Surveillance for the Monitoring of the COVID-19 Pandemic. Food Environ. Virol. 2021, doi:10.1007/s12560-021-09498-6.

2. Cluzel, N.;Courbariaux, M.;Wang, S.;Moulin, L.;Wurtzer, S.;Bertrand, I.;Laurent, K.;Monfort, P.;Gantzer, C.;Guyader, S.L.;et al. A Nationwide Indicator to Smooth and Normalize Heterogeneous SARS-CoV-2 RNA Data in Wastewater. Environ. Int. 2022, 158, 106998, doi:10.1016/j.envint.2021.106998.

3. Greenwald, H.D.;Kennedy, L.C.;Hinkle, A.;Whitney, O.N.;Fan, V.B.;Crits-Christoph, A.;Harris-Lovett, S.;Flamholz, A.I.;Al-Shayeb, B.;Liao, L.D.;et al. Tools for Interpretation of Wastewater SARS-CoV-2 Temporal and Spatial Trends Demonstrated with Data Collected in the San Francisco Bay Area. Water Res. X 2021, 12, 100111, doi:10.1016/j.wroa.2021.100111.

4. Ho, J.;Stange, C.;Suhrborg, R.;Wurzbacher, C.;Drewes, J.E.;Tiehm, A. SARS-CoV-2 Wastewater Surveillance in Germany: Long-Term PCR Monitoring, Suitability of Primer/Probe Combinations and Biomarker Stability; Epidemiology, 2021;

5. Diamond, M.B.;Keshaviah, A.;Bento, A.I.;Conroy-Ben, O.;Driver, E.M.;Ensor, K.B.;Halden, R.U.;Hopkins, L.P.;Kuhn, K.G.;Moe, C.L.;et al. Wastewater Surveillance of Pathogens Can Inform Public Health Responses. Nat. Med. 2022, 28, 1992–1995, doi:10.1038/s41591-022-01940-x.

6. Harris-Lovett, S.;Nelson, K.L.;Kantor, R.;Korfmacher, K.S. Wastewater Surveillance to Inform Public Health Decision Making in Residential Institutions. J. Public Health Manag. Pract. 2022, 10.1097/PHH.0000000000001636, doi:10.1097/PHH.0000000000001636.

7. Hopkins, L.;Persse, D.;Caton, K.;Ensor, K.;Schneider, R.;McCall, C.;Stadler, L.B. Citywide Wastewater SARS-CoV-2 Levels Strongly Correlated with Multiple Disease Surveillance Indicators and Outcomes over Three COVID-19 Waves. Sci. Total Environ. 2023, 855, 158967, doi:10.1016/j.scitotenv.2022.158967.

8. Usher, A.D. FIND Documents Dramatic Reduction in COVID-19 Testing. Lancet Infect. Dis. 2022, 22, 949, doi:10.1016/S1473-3099(22)00376-0.

9. Huisman, J.S.;Scire, J.;Caduff, L.;Fernandez, -Cassi Xavier; Ganesanandamoorthy, P.;Kull, A.;Scheidegger, A.;Stachler, E.;Boehm, A.B.;Hughes, B.;et al. Wastewater-Based Estimation of the Effective Reproductive Number of SARS-CoV-2. Environ. Health Perspect. 2022, 130, 057011, doi:10.1289/EHP10050.

10. Soller, J.;Jennings, W.;Schoen, M.;Boehm, A.;Wigginton, K.;Gonzalez, R.;Graham, K.E.;McBride, G.;Kirby, A.;Mattioli, M. Modeling Infection from SARS-CoV-2 Wastewater Concentrations: Promise, Limitations, and Future Directions. J. Water Health 2022, 20, 1197–1211, doi:10.2166/wh.2022.094.

11. McCall, C.;Fang, Z.N.;Li, D.;Czubai, A.J.;Juan, A.;LaTurner, Z.W.;Ensor, K.;Hopkins, L.;Bedient, P.B.;Stadler, L.B. Modeling SARS-CoV-2 RNA Degradation in Small and Large Sewersheds. Environ. Sci. Water Res. Technol. 2022, 8, 290–300, doi:10.1039/D1EW00717C.

12. Wade, M.J.;Lo Jacomo, A.;Armenise, E.;Brown, M.R.;Bunce, J.T.;Cameron, G.J.;Fang, Z.;Farkas, K.;Gilpin, D.F.;Graham, D.W.;et al. Understanding and Managing Uncertainty and Variability for Wastewater Monitoring beyond the Pandemic: Lessons Learned from the United Kingdom National COVID-19 Surveillance Programmes. J. Hazard. Mater. 2022, 424, 127456, doi:10.1016/j.jhazmat.2021.127456.

13. Bivins, A.;Greaves, J.;Fischer, R.;Yinda, K.C.;Ahmed, W.;Kitajima, M.;Munster, V.J.;Bibby, K. Persistence of SARS-CoV-2 in Water and Wastewater. Environ. Sci. Technol. Lett. 2020, 7, 937–942, doi:10.1021/acs.estlett.0c00730.

14. Farkas, K.;Adriaenssens, E.M.;Walker, D.I.;McDonald, J.E.;Malham, S.K.;Jones, D.L. Critical Evaluation of CrAssphage as a Molecular Marker for Human-Derived Wastewater Contamination in the Aquatic Environment. Food Environ. Virol. 2019, 11, 113–119, doi:10.1007/s12560-019-09369-1.

15. Langeveld, J.;Schilperoort, R.;Heijnen, L.;Elsinga, G.;Schapendonk, C.E.M.;Fanoy, E.;de Schepper, E.I.T.;Koopmans, M.P.G.;de Graaf, M.;Medema, G. Normalisation of SARS-CoV-2 Concentrations in Wastewater: The Use of Flow, Conductivity and CrAssphage; Epidemiology, 2021;

16. Hoar, C.;Li, Y.;Silverman, A.I. Assessment of Commonly Measured Wastewater Parameters to Estimate Sewershed Populations for Use in Wastewater-Based Epidemiology: Insights into Population Dynamics in New York City during the COVID-19 Pandemic. ACS EST Water 2022, acsestwater.2c00052, doi:10.1021/acsestwater.2c00052.

17. Yaniv, K.;Shagan, M.;Lewis, Y.E.;Kramarsky-Winter, E.;Weil, M.;Indenbaum, V.;Elul, M.;Erster, O.;Brown, A.S.;Mendelson, E.;et al. City-Level SARS-CoV-2 Sewage Surveillance. Chemosphere 2021, 283, 131194, doi:10.1016/j.chemosphere.2021.131194.

18. Carducci, A.;Federigi, I.;Liu, D.;Thompson, J.R.;Verani, M. Making Waves: Coronavirus Detection, Presence and Persistence in the Water Environment: State of the Art and Knowledge Needs for Public Health. Water Res. 2020, 179, 115907, doi:10.1016/j.watres.2020.115907.

19. CDC Wastewater Surveillance Testing Methods Available online: https://www.cdc.gov/healthywater/surveillance/wastewater-surveillance/testing-methods.html (accessed on 14 November 2022).

20. Noh, J.;Danuser, G. Estimation of the Fraction of COVID-19 Infected People in U.S. States and Countries Worldwide. PLOS ONE 2021, 16, e0246772. doi:10.1371/journal.pone.0246772.

21. World Health Organization Public Health Criteria to Adjust Public Health and Social Measures in the Context of COVID-19: Annex to Considerations in Adjusting Public Health and Social Measures in the Context of COVID-19; World Health Organization, 2020;

22. Chiu, W.A.;Ndeffo-Mbah, M.L. Using Test Positivity and Reported Case Rates to Estimate State-Level COVID-19 Prevalence and Seroprevalence in the United States. PLOS Comput. Biol. 2021, 17, e1009374. doi:10.1371/journal.pcbi.1009374.

23. Daza-Torres, M.L.;Montesinos-López, J.C.;Kim, M.;Olson, R.;Bess, C.W.;Rueda, L.;Susa, M.;Tucker, L.;García, Y.E.;Schmidt, A.J.;et al. Model Training Periods Impact Estimation of COVID-19 Incidence from Wastewater Viral Loads. Sci. Total Environ. 2022, 159680, doi:10.1016/j.scitotenv.2022.159680.

24. Feng, S.;Roguet, A.;McClary-Gutierrez, J.S.;Newton, R.J.;Kloczko, N.;Meiman, J.G.;McLellan, S.L. Evaluation of Sampling, Analysis, and Normalization Methods for SARS-CoV-2 Concentrations in Wastewater to Assess COVID-19 Burdens in Wisconsin Communities. ACS EST Water 2021, 1, 1955–1965, doi:10.1021/acsestwater.1c00160.

25. Maal-Bared, R.;Qiu, Y.;Li, Q.;Gao, T.;Hrudey, S.E.;Bhavanam, S.;Ruecker, N.J.;Ellehoj, E.;Lee, B.E.;Pang, X. Does Normalization of SARS-CoV-2 Concentrations by Pepper Mild Mottle Virus Improve Correlations and Lead Time between Wastewater Surveillance and Clinical Data in Alberta (Canada): Comparing Twelve SARS-CoV-2 Normalization Approaches. Sci. Total Environ. 2023, 856, 158964, doi:10.1016/j.scitotenv.2022.158964.

26. Mitranescu, A.;Uchaikina, A.;Kau, A.-S.;Stange, C.;Ho, J.;Tiehm, A.;Wurzbacher, C.;Drewes, J.E. Wastewater-Based Epidemiology for SARS-CoV-2 Biomarkers: Evaluation of Normalization Methods in Small and Large Communities in Southern Germany. ACS EST Water 2022, doi:10.1021/acsestwater.2c00306.

27. Hamed, K.H. Effect of Persistence on the Significance of Kendall’s Tau as a Measure of Correlation between Natural Time Series. Eur. Phys. J. Spec. Top. 2009, 174, 65–79, doi:10.1140/epjst/e2009-01090-x.

28. Wolfe, M.K.;Archana, A.;Catoe, D.;Coffman, M.M.;Dorevich, S.;Graham, K.E.;Kim, S.;Grijalva, L.M.;Roldan-Hernandez, L.;Silverman, A.I.;et al. Scaling of SARS-CoV-2 RNA in Settled Solids from Multiple Wastewater Treatment Plants to Compare Incidence Rates of Laboratory-Confirmed COVID-19 in Their Sewersheds. Environ. Sci. Technol. Lett. 2021, 8, 398–404, doi:10.1021/acs.estlett.1c00184.

29. Nourbakhsh, S.;Fazil, A.;Li, M.;Mangat, C.S.;Peterson, S.W.;Daigle, J.;Langner, S.;Shurgold, J.;D’Aoust, P.;Delatolla, R.;et al. A Wastewater-Based Epidemic Model for SARS-CoV-2 with Application to Three Canadian Cities. Epidemics 2022, 39, 100560, doi:10.1016/j.epidem.2022.100560.

30. Rodríguez Rasero, F.J.;Moya Ruano, L.A.;Rasero Del Real, P.;Cuberos Gómez, L.;Lorusso, N. Associations between SARS-CoV-2 RNA Concentrations in Wastewater and COVID-19 Rates in Days after Sampling in Small Urban Areas of Seville: A Time Series Study. Sci. Total Environ. 2022, 806, 150573, doi:10.1016/j.scitotenv.2021.150573.

31. Zdenkova, K.;Bartackova, J.;Cermakova, E.;Demnerova, K.;Dostalkova, A.;Janda, V.;Jarkovsky, J.;Lopez Marin, M.A.;Novakova, Z.;Rumlova, M.;et al. Monitoring COVID-19 Spread in Prague Local Neighborhoods Based on the Presence of SARS-CoV-2 RNA in Wastewater Collected throughout the Sewer Network. Water Res. 2022, 216, 118343, doi:10.1016/j.watres.2022.118343.

32. Kantor, R.S.;Greenwald, H.D.;Kennedy, L.C.;Hinkle, A.;Harris-Lovett, S.;Metzger, M.;Thornton, M.M.;Paluba, J.M.;Nelson, K.L. Operationalizing a Routine Wastewater Monitoring Laboratory for SARS-CoV-2. PLOS Water 2022, 1, e0000007. doi:10.1371/journal.pwat.0000007.

33. Whitney, O.N.;Kennedy, L.C.;Fan, V.B.;Hinkle, A.;Kantor, R.;Greenwald, H.;Crits-Christoph, A.;Al-Shayeb, B.;Chaplin, M.;Maurer, A.C.;et al. Sewage, Salt, Silica, and SARS-CoV-2 (4S): An Economical Kit-Free Method for Direct Capture of SARS-CoV-2 RNA from Wastewater. Environ. Sci. Technol. 2021, 55, 4880–4888, doi:10.1021/acs.est.0c08129.

34. Menne, M.J.;Durre, I.;Korzeniewski, B.;McNeill, S.;Thomas, K.;Yin, X.;Anthony, S.;Ray, R.;Vose, R.S.;Gleason, B.E.;et al. Global Historical Climatology Network - Daily (GHCN-Daily), Version 3 2012.

35. COVID-19 Time-Series Metrics by County and State 2022.

36. CDPH COVID-19 Variant Data Available online: https://data.chhs.ca.gov/dataset/covid-19-variant-data (accessed on 17 November 2022).

37. COVID CG Available online: https://covidcg.org/ (accessed on 22 November 2022).

38. Zheng, X.;Li, S.;Deng, Y.;Xu, X.;Ding, J.;Lau, F.T.K.;In Yau, C.;Poon, L.L.M.;Tun, H.M.;Zhang, T. Quantification of SARS-CoV-2 RNA in Wastewater Treatment Plants Mirrors the Pandemic Trend in Hong Kong. Sci. Total Environ. 2022, 844, 157121, doi:10.1016/j.scitotenv.2022.157121.

39. Graham, K.E.;Loeb, S.K.;Wolfe, M.K.;Catoe, D.;Sinnott-Armstrong, N.;Kim, S.;Yamahara, K.M.;Sassoubre, L.M.;Mendoza Grijalva, L.M.;Roldan-Hernandez, L.;et al. SARS-CoV-2 RNA in Wastewater Settled Solids Is Associated with COVID-19 Cases in a Large Urban Sewershed. Environ. Sci. Technol. 2021, 55, 488–498, doi:10.1021/acs.est.0c06191.

40. Kim, S.;Kennedy, L.C.;Wolfe, M.K.;Criddle, C.S.;Duong, D.H.;Topol, A.;White, B.J.;Kantor, R.S.;Nelson, K.L.;Steele, J.A.;et al. SARS-CoV-2 RNA Is Enriched by Orders of Magnitude in Solid Relative to Liquid Wastewater at Publicly Owned Treatment Works; Infectious Diseases (except HIV/AIDS), 2021;

41. Nagarkar, M.;Keely, S.P.;Jahne, M.;Wheaton, E.;Hart, C.;Smith, B.;Garland, J.;Varughese, E.A.;Braam, A.;Wiechman, B.;et al. SARS-CoV-2 Monitoring at Three Sewersheds of Different Scales and Complexity Demonstrates Distinctive Relationships between Wastewater Measurements and COVID-19 Case Data. Sci. Total Environ. 2021, 151534, doi:10.1016/j.scitotenv.2021.151534.

42. Gudra, D.;Dejus, S.;Bartkevics, V.;Roga, A.;Kalnina, I.;Strods, M.;Rayan, A.;Kokina, K.;Zajakina, A.;Dumpis, U.;et al. Detection of SARS-CoV-2 RNA in Wastewater and Importance of Population Size Assessment in Smaller Cities: An Exploratory Case Study from Two Municipalities in Latvia. Sci. Total Environ. 2022, 823, 153775, doi:10.1016/j.scitotenv.2022.153775.

43. Centers for Disease Control and Prevention Developing a Wastewater Surveillance Sampling Strategy 2022.

44. Hill, D.T.;Cousins, H.;Dandaraw, B.;Faruolo, C.;Godinez, A.;Run, S.;Smith, S.;Willkens, M.;Zirath, S.;Larsen, D.A. Wastewater Treatment Plant Operators Report High Capacity to Support Wastewater Surveillance for COVID-19 across New York State, USA. Sci. Total Environ. 2022, 837, 155664, doi:10.1016/j.scitotenv.2022.155664.

45. Lieberman-Cribbin, W.;Tuminello, S.;Flores, R.M.;Taioli, E. Disparities in COVID-19 Testing and Positivity in New York City. Am. J. Prev. Med. 2020, 59, 326–332, doi:10.1016/j.amepre.2020.06.005.

46. Vaughan, L.;Zhang, M.;Gu, H.;Rose, J.B.;Naughton, C.C.;Medema, G.;Allan, V.;Roiko, A.;Blackall, L.;Zamyadi, A. An Exploration of Challenges Associated with Machine Learning for Time Series Forecasting of COVID-19 Community Spread Using Wastewater-Based Epidemiological Data. Sci. Total Environ. 2023, 858, 159748, doi:10.1016/j.scitotenv.2022.159748.

47. Medina, C.Y.;Kadonsky, K.F.;Roman, F.A.;Tariqi, A.Q.;Sinclair, R.G.;D’Aoust, P.M.;Delatolla, R.;Bischel, H.N.;Naughton, C.C. The Need of an Environmental Justice Approach for Wastewater Based Epidemiology for Rural and Disadvantaged Communities: A Review in California. Curr. Opin. Environ. Sci. Health 2022, 27, 100348, doi:10.1016/j.coesh.2022.100348.

48. Olesen, S.W.;Imakaev, M.;Duvallet, C. Making Waves: Defining the Lead Time of Wastewater-Based Epidemiology for COVID-19. Water Res. 2021, 202, 117433, doi:10.1016/j.watres.2021.117433.

49. Bibby, K.;Bivins, A.;Wu, Z.;North, D. Making Waves: Plausible Lead Time for Wastewater Based Epidemiology as an Early Warning System for COVID-19. Water Res. 2021, 202, 117438, doi:10.1016/j.watres.2021.117438.

50. Xiao, A.;Wu, F.;Bushman, M.;Zhang, J.;Imakaev, M.;Chai, P.R.;Duvallet, C.;Endo, N.;Erickson, T.B.;Armas, F.;et al. Metrics to Relate COVID-19 Wastewater Data to Clinical Testing Dynamics. Water Res. 2022, 118070, doi:10.1016/j.watres.2022.118070.

51. Li, B.;Deng, A.;Li, K.;Hu, Y.;Li, Z.;Shi, Y.;Xiong, Q.;Liu, Z.;Guo, Q.;Zou, L.;et al. Viral Infection and Transmission in a Large, Well-Traced Outbreak Caused by the SARS-CoV-2 Delta Variant. Nat. Commun. 2022, 13, 460, doi:10.1038/s41467-022-28089-y.

52. Prasek, S.M.;Pepper, I.L.;Innes, G.K.;Slinski, S.;Ruedas, M.;Sanchez, A.;Brierley, P.;Betancourt, W.Q.;Stark, E.R.;Foster, A.R.;et al. Population Level SARS-CoV-2 Fecal Shedding Rates Determined via Wastewater-Based Epidemiology. Sci. Total Environ. 2022, 838, 156535, doi:10.1016/j.scitotenv.2022.156535.

53. Bloemen, M.;Delang, L.;Rector, A.;Raymenants, J.;Thibaut, J.;Pussig, B.;Fondu, L.;Aertgeerts, B.;Van Ranst, M.;Van Geet, C.;et al. Detection Of SARS-COV-2 Variants Of Concern In Wastewater Of Leuven, Belgium; Epidemiology, 2022;

54. Yuan, S.;Ye, Z.-W.;Liang, R.;Tang, K.;Zhang, A.J.;Lu, G.;Ong, C.P.;Poon, V.K.-M.;Chan, C.C.-S.;Mok, B.W.-Y.;et al. The SARS-CoV-2 Omicron (B.1.1.529) Variant Exhibits Altered Pathogenicity, Transmissibility, and Fitness in the Golden Syrian Hamster Model; Microbiology, 2022;

55. Bramante, C.T.;Proper, J.L.;Boulware, D.R.;Karger, A.B.;Murray, T.;Rao, V.;Hagen, A.;Tignanelli, C.J.;Puskarich, M.;Cohen, K.;et al. Vaccination Against SARS-CoV-2 Is Associated With a Lower Viral Load and Likelihood of Systemic Symptoms. Open Forum Infect. Dis. 2022, 9, ofac066. doi:10.1093/ofid/ofac066.

56. Levine-Tiefenbrun, M.;Yelin, I.;Katz, R.;Herzel, E.;Golan, Z.;Schreiber, L.;Wolf, T.;Nadler, V.;Ben-Tov, A.;Kuint, J.;et al. Initial Report of Decreased SARS-CoV-2 Viral Load after Inoculation with the BNT162b2 Vaccine. Nat. Med. 2021, 27, 790–792, doi:10.1038/s41591-021-01316-7.

57. McEllistrem, M.C.;Clancy, C.J.;Buehrle, D.J.;Lucas, A.;Decker, B.K. Single Dose of an MRNA Severe Acute Respiratory Syndrome Coronavirus 2 (SARS-Cov-2) Vaccine Is Associated With Lower Nasopharyngeal Viral Load Among Nursing Home Residents With Asymptomatic Coronavirus Disease 2019 (COVID-19). Clin. Infect. Dis. 2021, 73, e1365–e1367, doi:10.1093/cid/ciab263.

58. Zhang, N.;Gong, Y.;Meng, F.;Shi, Y.;Wang, J.;Mao, P.;Chuai, X.;Bi, Y.;Yang, P.;Wang, F. Comparative Study on Virus Shedding Patterns in Nasopharyngeal and Fecal Specimens of COVID-19 Patients. Sci. China Life Sci. 2021, 64, 486–488, doi:10.1007/s11427-020-1783-9.

