## Supplementary figures for "The dynamic relationship between COVID-19 cases and SARS-CoV-2 wastewater concentrations across time and space: considerations for model training data sets"

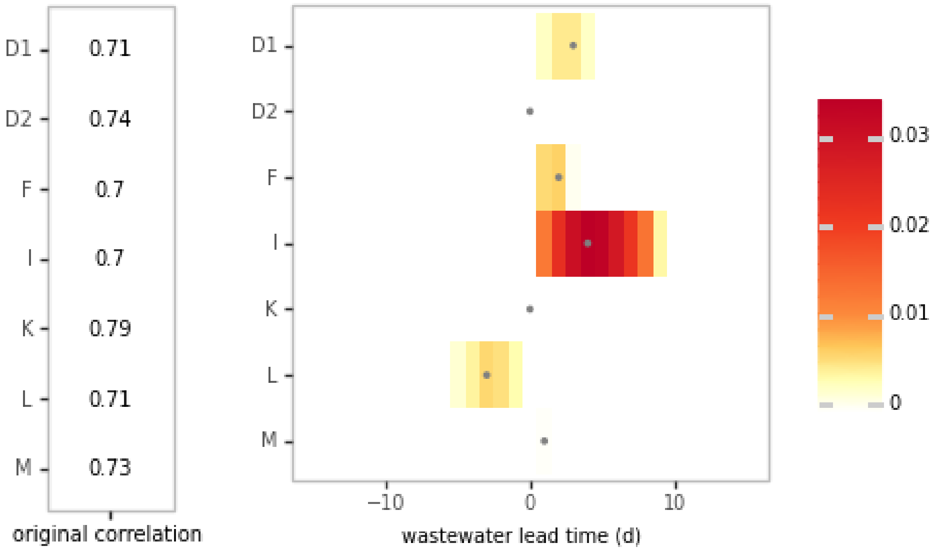


**Figure S1A.** Kendall's correlations between wastewater SARS-CoV-2 RNA and COVID-19 cases per 100,000 people without timeshift (left) and cross-correlation heatmap of changes in Kendall's tau after shifts of up to -14 to +14 days were applied to case data (right). The maximum correlation is indicated by gray points. Prior to correlation calculations, all wastewater data were flow-normalized, log-scaled, and lowess smoothed (with interpolation) to reduce noise, and all case data were converted to 7-day moving averages and log-scaled. Sewersheds were included only if they were sampled throughout the entire time series.

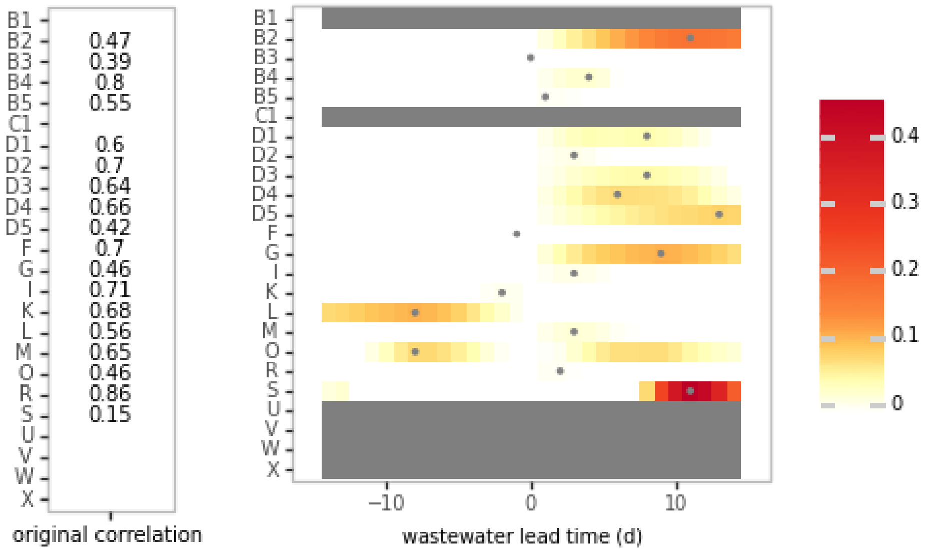


**Figure S1B.** Kendall's correlations between wastewater SARS-CoV-2 RNA and COVID-19 cases per 100,000 people during the Epsilon/Alpha variant-dominated surge without time shift (left) and after shifts of up to -14 to +14 days were applied to case data, (right), as in Figure S1A.

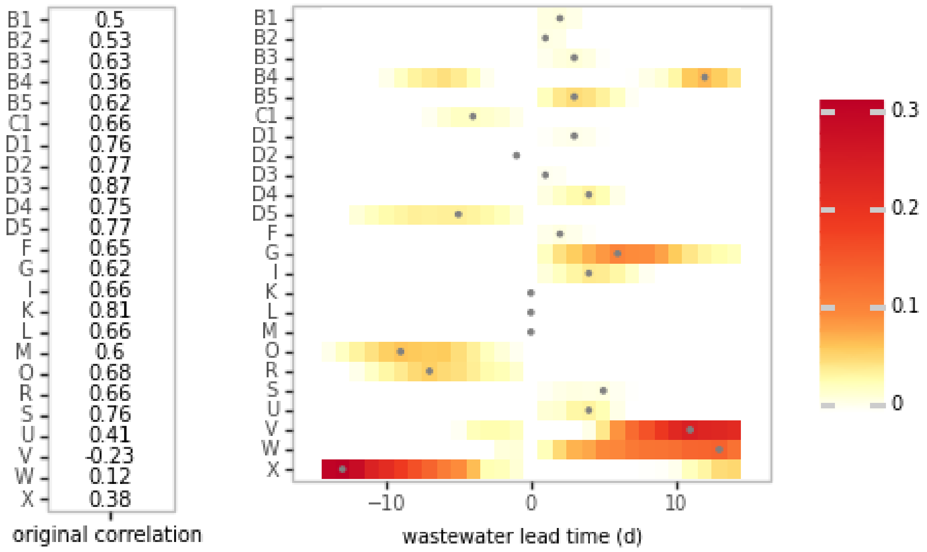


**Figure S1C.** Kendall's correlations between wastewater SARS-CoV-2 RNA and COVID-19 cases per 100,000 people during the Delta variant-dominated surge without time shift (left) and after shifts of up to -14 to +14 days were applied to case data, (right), as in Figure S1A.

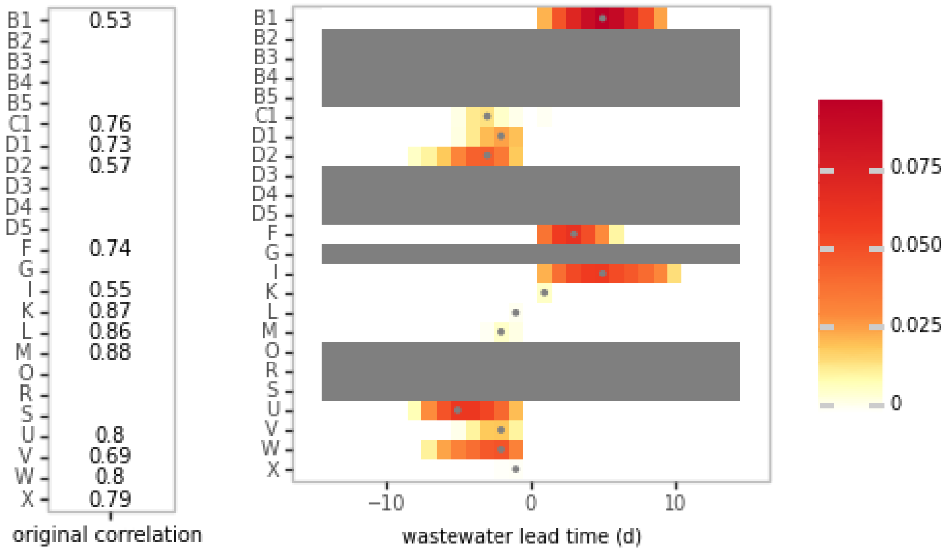


**Figure S1D.** Kendall's correlations between wastewater SARS-CoV-2 RNA and COVID-19 cases per 100,000 people during the Omicron BA.1 variant-dominated surge without time shift (left) and after shifts of up to -14 to +14 days were applied to case data, (right), as in Figure S1A.

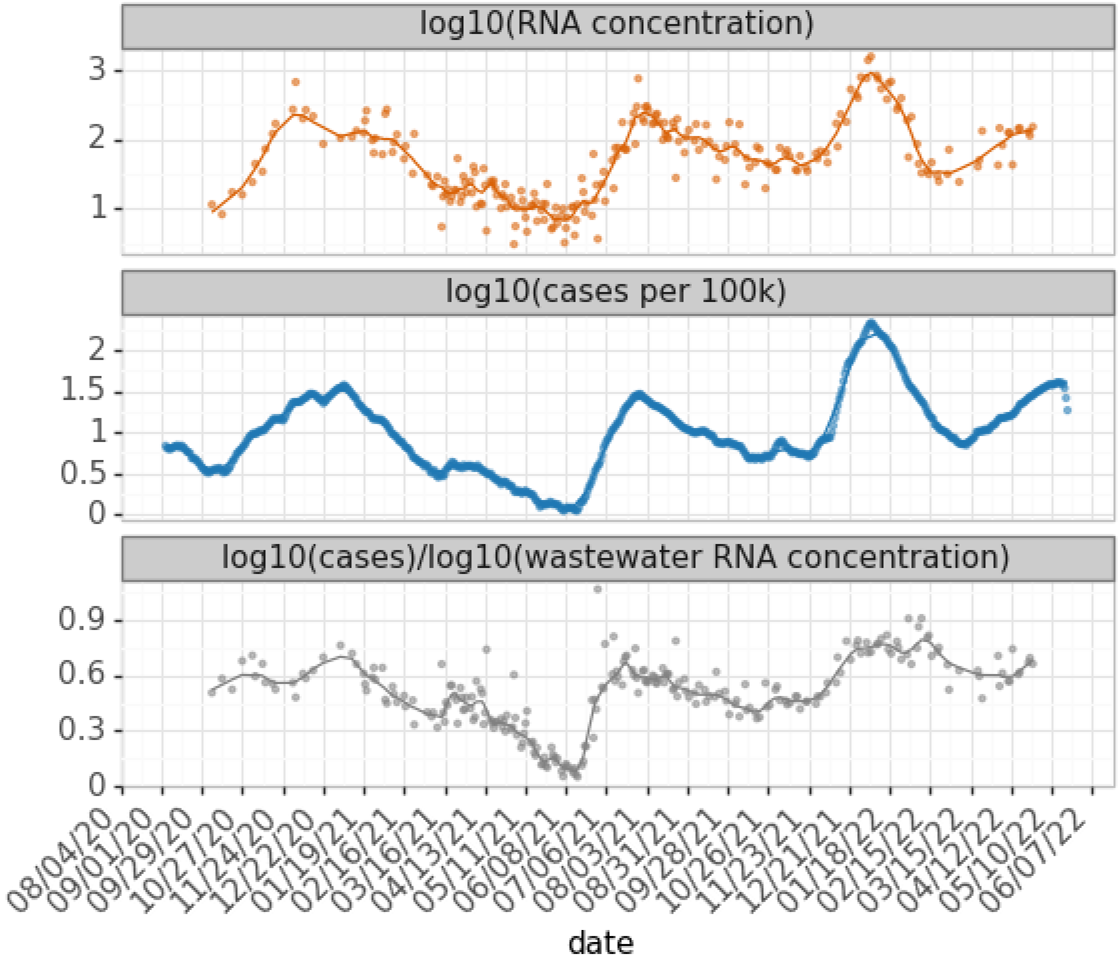


**Figure S2.** Time series of flow-normalized wastewater SARS-CoV-2 RNA concentration (gc/mL, orange), 7-day moving average COVID-19 cases per 100,000 people (blue), the ratio of log-scaled cases over log-scaled RNA concentration (gray), and lowess-smoothed curves (alpha = 0.05), at site D1 from August 25, 2020 to May 31, 2022.

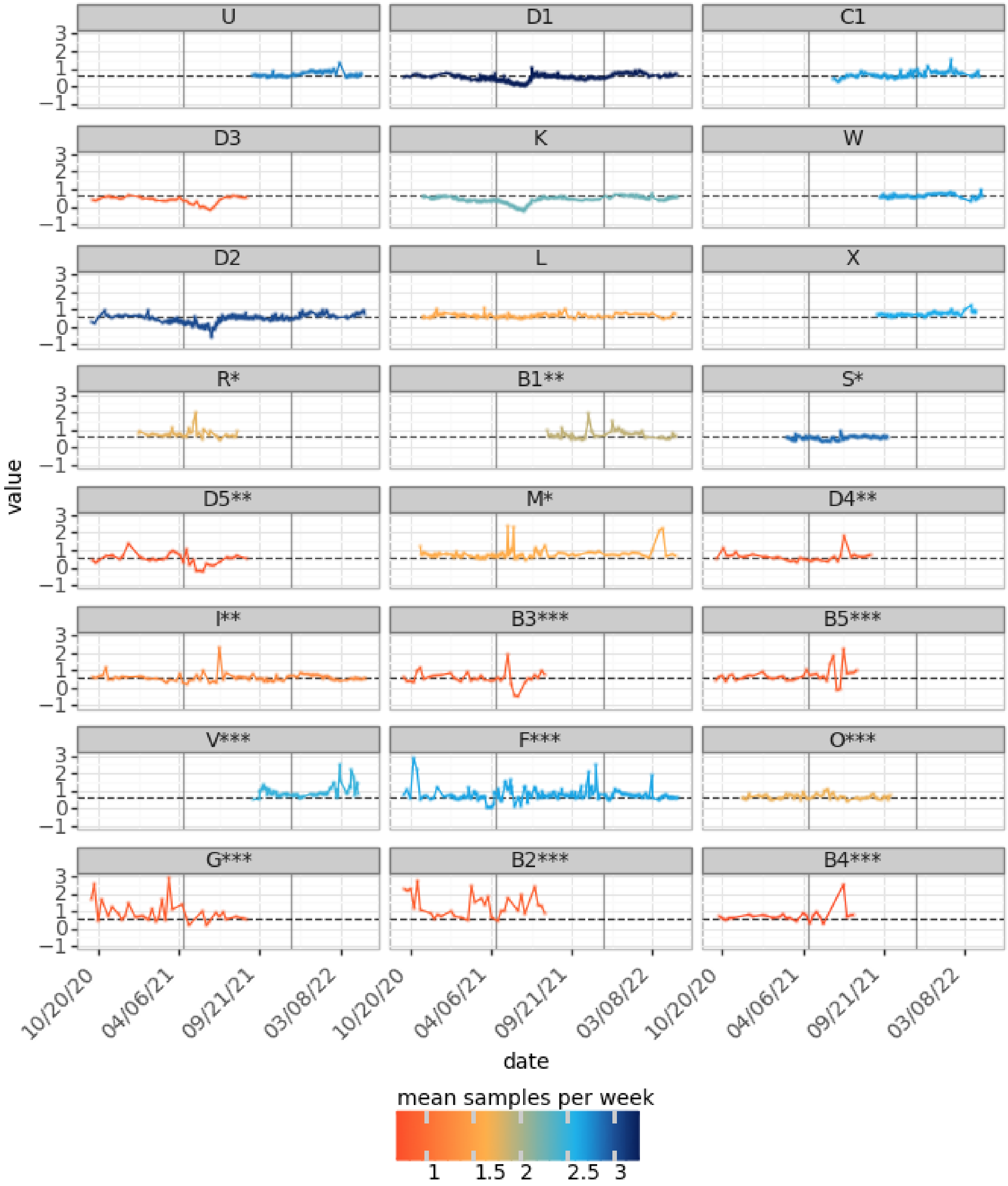


**Figure S3.** Time series of the ratio between log10(COVID-19 cases) and log10(wastewater SARS-CoV-2 RNA concentration) at 24 sites. Graphs are sorted by sewershed population and colored by mean weekly sampling frequency. Fraction of case data that is masked is represented by the following: *0-0.05 **0.05-0.33 ***>0.33. One outlier at site F was removed for visualization.

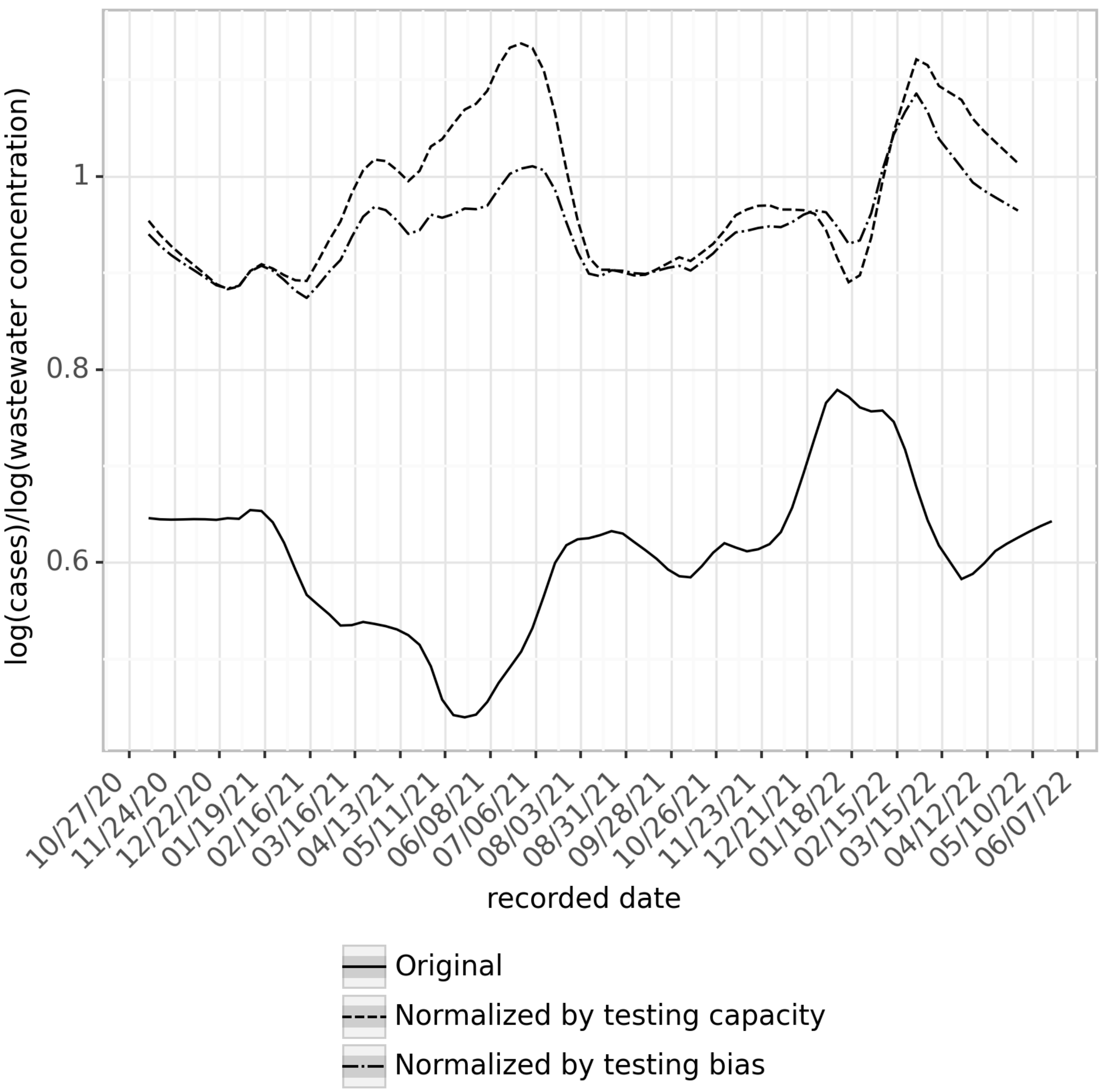


**Figure S4**. Lowess-smoothed (alpha=0.1) ratio of log(cases) to log(wastewater concentration) non-normalized, normalized by testing capacity (Eq. 4) and normalized by testing bias (Eq. 5) at 24 sewersheds. Averages were calculated for each week and each sewershed. For each sewershed, for a given week, a minimum of two data points was required for inclusion.

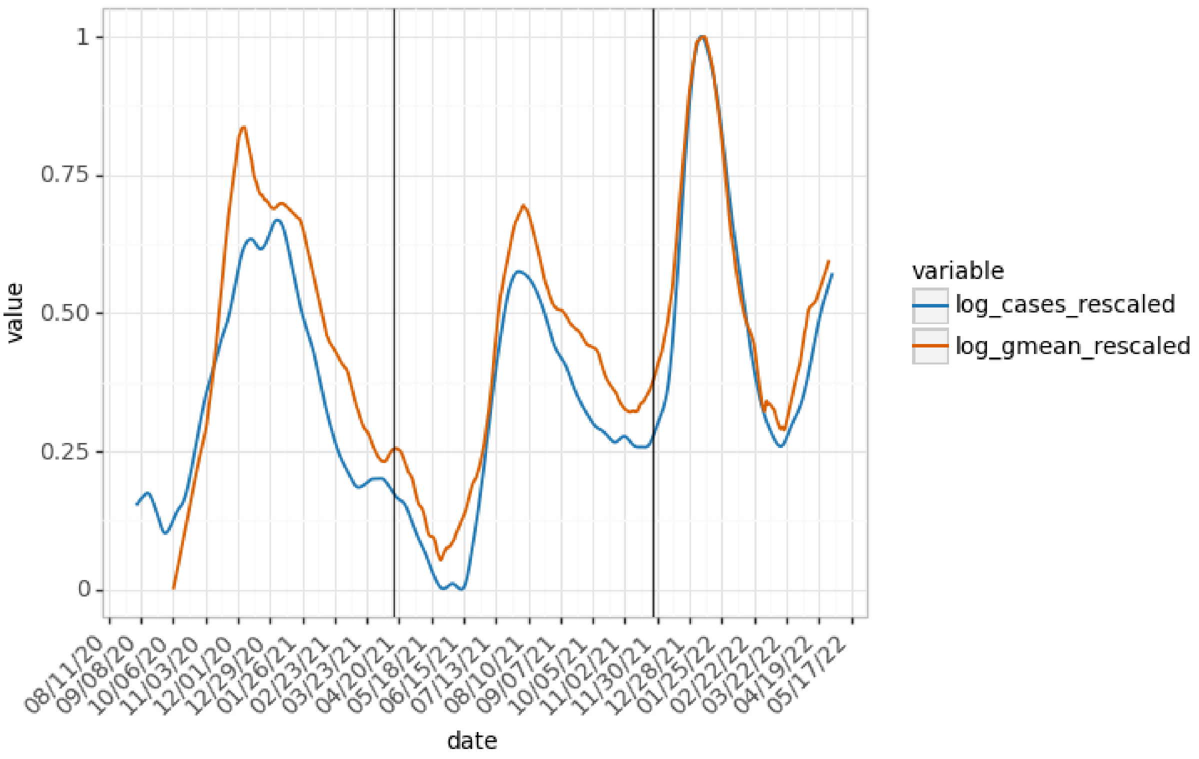


**Figure S5.** Lowess smoothed time series of log-scaled, flow-normalized wastewater SARS-CoV-2 RNA concentration (gc/mL, orange) and log-scaled, 7-day moving average COVID-19 cases per 100,000 people (blue) at sites D1, D2, K, L, and M. Both time series were rescaled by defining the minimum as 0 and the maximum as 1.
